# Bridging the Genomic Equity Gap with Context-Enhanced Risk Stratification in American Indians: the Strong Heart Study

**DOI:** 10.64898/2026.02.08.26345859

**Authors:** Jiawen Du, Andrea R.V.R. Horimoto, Lyle G. Best, Ying Zhang, Shelley A. Cole, Jason G. Umans, Nora Franceschini, Quan Sun

## Abstract

Polygenic scores (PGS) show promise for disease risk stratification but suffer from limited portability across populations. American Indians face a disproportionate burden of cardiovascular disease yet remain significantly underrepresented in genomic research, limiting equitable access to precision medicine. Here, we evaluate whether integrating specific lifestyle and clinical context variables with PGS enhances risk prediction for cardiometabolic traits in 424,622 European from UK Biobank (UKB) and 3,157 American Indian populations from the Strong Heart Study (SHS). By comparing genetics-only models to full models incorporating context variables and gene-context interactions across blood pressure traits, coronary heart disease (CHD), and stroke, we found that the integration of context variables significantly improved prediction accuracy in both cohorts. Notably, for American Indian participants, the new model incorporating context and genetic interactions significantly improved model discrimination for CHD compared to an established clinical risk model. These findings suggest that modeling the interplay between inherited risk and modifiable factors can recover predictive power loss due to imperfect PGS transferability, offering a viable pathway toward more equitable and effective precision medicine for under-represented populations.

## Introduction

Polygenic scores (PGSs), which aggregate the effects of genetic variants into a single metric of inherited risk, have shown success in identifying individuals at high risk for a range of cardiometabolic diseases^1–8^. It often outperforms traditional clinical risk factors alone^2,9^, demonstrating potential clinical promise^10,11^. However, the clinical utility of PGSs is hampered by their development in predominantly European (EUR) ancestry populations^12^, leading to attenuated performance and biased risk estimates in other populations. There have been widespread calls and efforts to include diverse populations in genomic research to mitigate health disparities^12–15^, with a wide range of PGS methods developed specifically for non-European populations^8,16–19^, yet many populations remain underrepresented in genome-wide association studies (GWAS)^13,20^. American Indian individuals, with a substantially higher burden of cardiovascular diseases, are among the most underrepresented in genomic research, resulting in a gap in equitable precision medicine^7,8,21^.

Recent studies have shown that PGSs show context specificity, where PGS performance may vary across different levels of demographic, lifestyle, and clinical variables, such as age, sex, smoking status, lipid levels, among others^3,22^, indicating joint influence between PGS and context variables. Some studies treat these context variables as environmental factors, and have explored the interactions between PGS and these broadly-defined environmental variables in some diseases^23–26^. While these context variables have been utilized in risk assessment as clinical risk factors for American Indian populations^27^, there has not been any attempt to exploring their interplay with PGS in this population group. Direct comparisons of these gene-environment interaction (GxE) or gene-context interaction (GxC) effects between well-studied European populations and American Indians are also not yet available to the best of our knowledge. Additionally, it remains unclear whether incorporating GxC effects will mitigate the transferability of genetic prediction models across populations.

In this study, we bridge this gap by leveraging two distinct cohorts: UK Biobank (UKB), a large, well-studied population of primarily European ancestry; and Strong Heart Study (SHS), an American Indian cohort. Focusing on cardiovascular traits, we first investigate which key context factors can enhance risk prediction in addition to PGS. Second, we quantify the improvement in risk prediction gained by incorporating these context factors and their interactions with genetic effects (i.e. PGS). Our study demonstrates that integrating specific context variables and their interactions with PGS yields consistent and significant improvements in risk prediction for blood pressure (BP) and coronary heart disease (CHD) across both European and American Indian populations, while highlighting distinct population-specific risk profiles.

## Results

### Study Overview

In this study, we focused on studying the interactions between PGS and clinical risk factors (as context variables) for five cardiovascular traits including systolic, diastolic, and pulse blood pressure (SBP, DBP, and PP), coronary heart disease (CHD), and stroke, in diverse populations, leveraging data of 424,622 European ancestry individuals from UKB and 3,157 American Indian participants from SHS (**Figure 1**). We first evaluated the contribution of each single context variable to trait prediction while accounting for PGS, separately in each population, to identify the shared and distinct GxC patterns by comparing a baseline genetics-only model against a comprehensive GxC model that incorporated context factors and their interactions with PGS (**Methods**). We further built prediction models including all the identified context variables and their interactions with PGS, and quantified the improvement in predictive accuracy in both UKB and SHS, compared to genetics-only models. Finally, focusing on American Indians, we compared our new models with previously established clinical risk prediction models developed specifically for this community^28^, to show that our models, by integrating PGS and clinical risk factors, have the potential to become the new standard in evaluating American Indian cardiovascular health.

**Figure 1.**
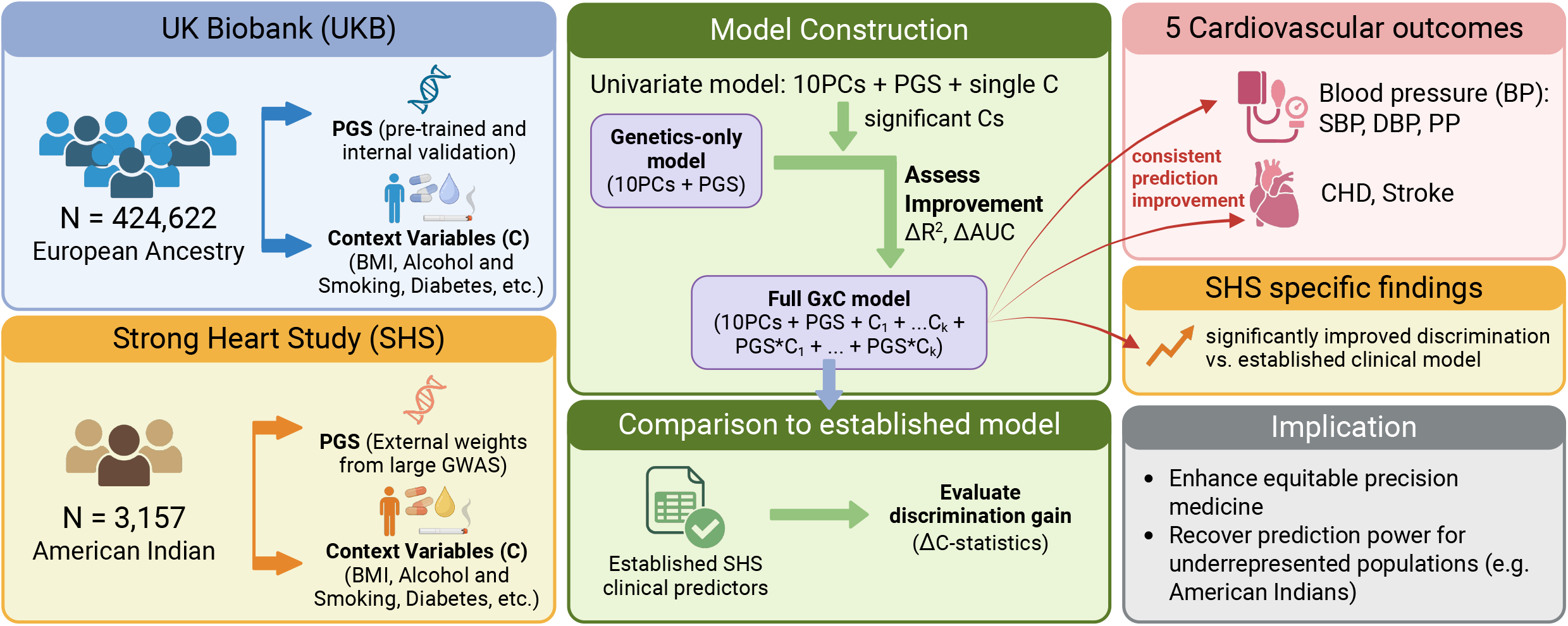
Study Overview. Our study integrated PGS with lifestyle and clinical context variables to enhance cardiometabolic risk prediction in diverse populations, specifically comparing 424,622 European ancestry individuals from the UKB and 3,157 American Indian participants from the SHS. We evaluated five cardiovascular traits, systolic, diastolic, and pulse blood pressure (SBP, DBP, and PP), coronary heart disease (CHD), and stroke, by comparing a baseline genetics-only model against a comprehensive GxC model that incorporated context factors and their interactions with PGS. To assess clinical utility, we quantified improvements in predictive accuracy through changes in partial R2 and Area Under the Curve (AUC) across both cohorts and further benchmarked the GxC model’s performance against an established clinical risk prediction model for American Indians using Harrell’s C-index.

### Characteristics of UKB and SHS cohorts

For SHS, we incorporated 3,157 individuals after QC (**Methods**), with 58.4% females (N = 1,845). The mean age at enrollment was 56.4 years, ranging from 44.5 to 75.4 years. Baseline mean SBP and DBP were 126.5 mmHg and 76.4 mmHg, resulting in a mean PP to be 50.0 mmHg. Antihypertensive medication was reported by 22.6% of participants. After adjustment for medication use, the mean SBP, DBP, and PP increased to 129.9, 78.8 and 51.2. The prevalence of CHD and stroke were 38.3% and 11.4%, respectively.

The UKB analysis included 424,622 individuals of European ancestry (**Methods**). Their mean age was 67.7 years and 54.2% were female. The baseline average SBP, DBP and PP measurements were 138.0 mmHg, 82.2 mmHg and 55.8 mmHg. Within this group, 20.6% of them reported using antihypertensive medication. After correction, these mean values increased to 141.1, 84.3, and 56.9 mmHg. The summary statistics of all baseline characteristics and context variables for both cohorts are listed in **Table 1**.

**Table 1.**
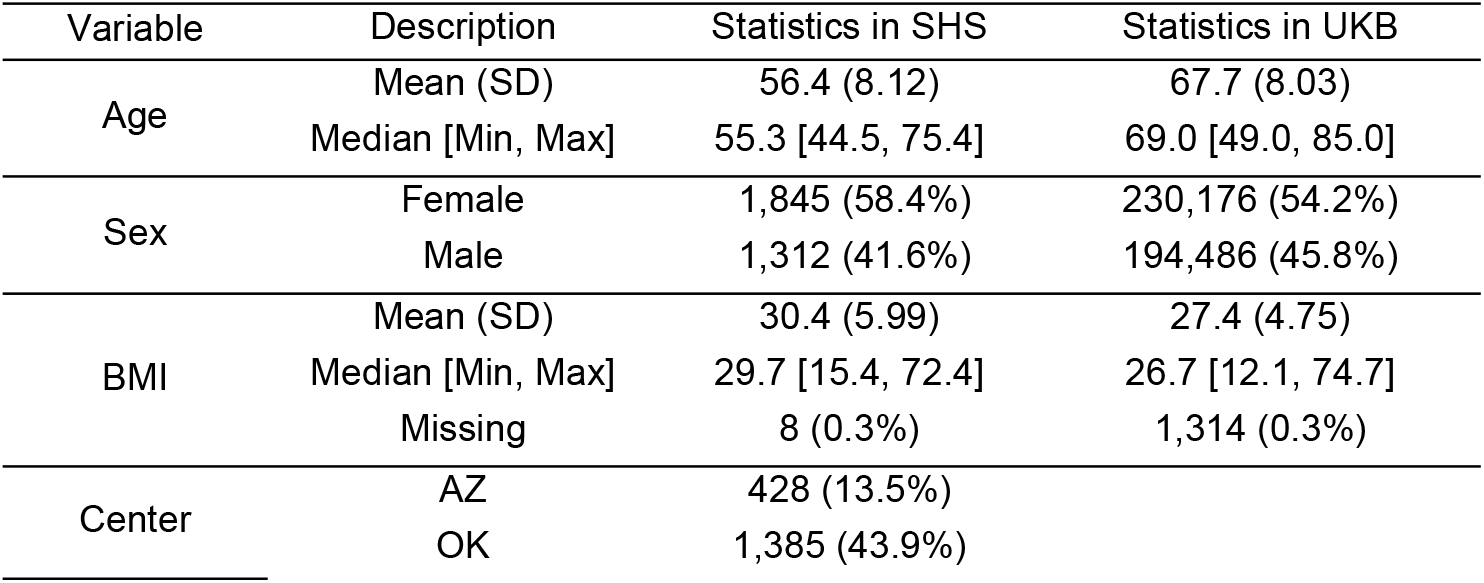
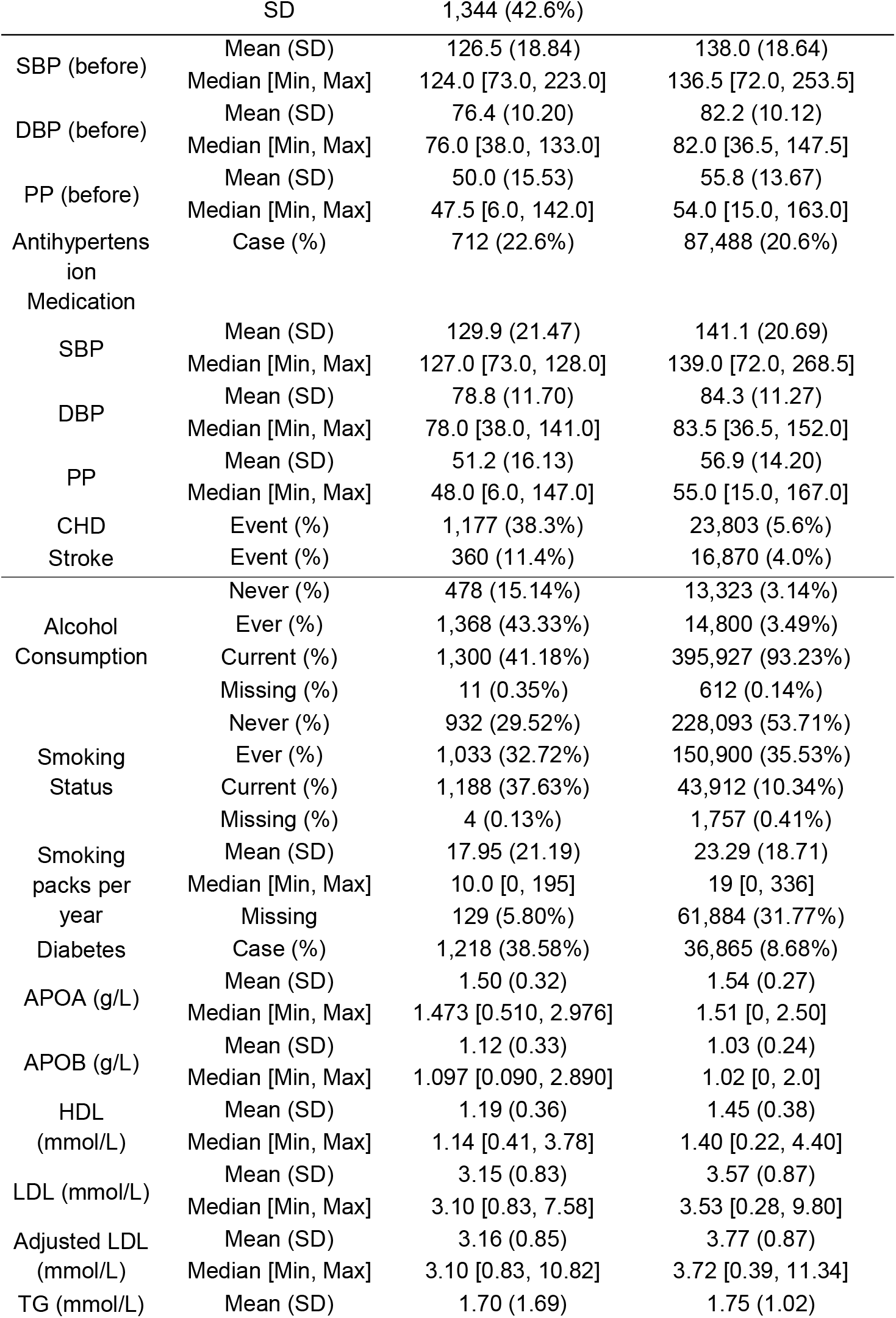
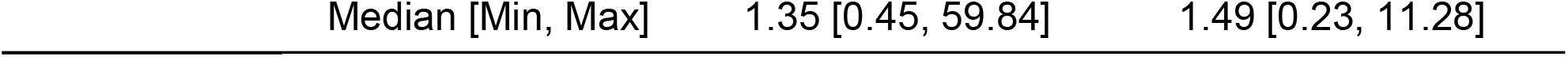
Characteristics of study participants in SHS and UKB.

### Identification of contributing context variables

We first screened context variables in a series of univariate GxC models to identify context and GxC terms that contribute to BP values and CHD and stroke risks (**Methods**). For BP traits, we identified several shared patterns between the two cohorts (**Figure 2, Supplementary Tables 1-2, Supplementary Figures 1-2**). For example, current smoking was significantly associated with lower SBP and DBP (**Figure 2a**), where DBP showed greater impact in SHS (effect size = - 2.58, p-value = 1.0E-5) than in UKB (effect size = -0.68, p-value = 3.4E-5). This finding is consistent with the association between higher smoking pack-years and lower DBP in both cohorts. In addition, diabetes was associated with a significant increase in SBP and PP where SBP showed greater impact by diabetes in SHS (effect size = 4.46, p-value = 9.3E-7) than in UKB (effect size = 1.99, p-value = 5.8E-11). Lipids, while most of them served as significant context variables, also demonstrated consistency between the two cohorts. Specifically, higher levels of APOA and HDL were positively associated with all the three BP traits in both cohorts; while APOB and TG showed significant positive associations in both cohorts with SBP and DBP.

**Figure 2.**
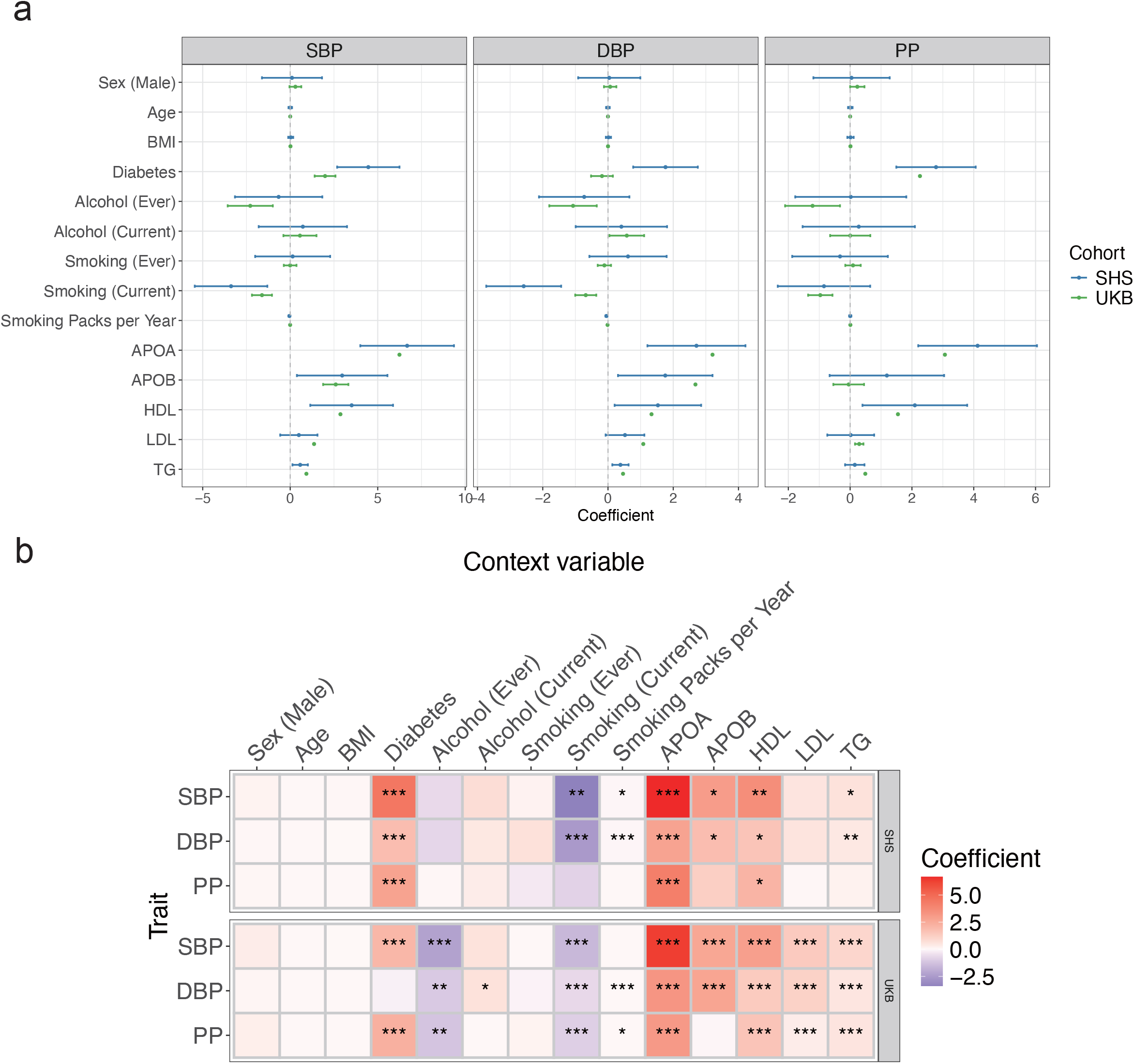
Univariate tests of association between blood pressure traits and context variables. **a**. Forest plots of coefficients of context variables for systolic blood pressure (SBP), diastolic blood pressure (DBP), and pulse pressure (PP) in SHS (blue lines) and UKB (green lines). The x-axis represents the effect size of each context variable and error bars indicate 95% confidence intervals. **b**. Heatmap of coefficients of context variables for SBP, DBP and PP in SHS (upper panel) and UKB (lower panel). Asterisks represent significance (p-value < 0.05(*)/0.01(**)/0.001(***)).

We also noticed several cohort-specific patterns. For example, ever consuming alcohol showed a negative association with the three BP traits in UKB (effect size = -2.28, -1.07, -1.22, p-value = 5.2E-4, 3.9E-3, 7.3E-3 for SBP, DBP and PP, respectively) while none of them was significant in SHS (effect size = -0.66, -0.73, 0.02, p-value = 0.60, 0.30, 0.98 for SBP, DBP and PP, respectively). We note that 93.2% UKB participants reported current drinking, while only 41.2% SHS participants were currently consuming alcohol, which potentially explained the difference in the association results here.

For cardiovascular outcomes (i.e., CHD and stroke), we identified diabetes status as a significant risk factor and HDL as a significant protective factor consistent in both outcomes and in both cohorts (**Figure 3a-b, Supplementary Tables 3-4, Supplementary Figures 3-4**). In addition, ever drinking and ever smoking showed consistent significant associations with higher disease risk; while for stroke, HDL showed consistent protective association in both cohorts. However, we also observed inconsistent associations between the two cohorts. While APOB was significantly associated with CHD in both populations, their effects were discordant where it increased risk in SHS but appeared protective in UKB (**Figure 3c**). We consider this counter-intuitive finding in UKB is likely attributable to confounding by medication, where individuals with high cardiovascular risks are treated with lipid-lowering therapies, and artificially lowering their lipid levels while their risk remains elevated. This hypothesis was confirmed by the inconsistencies of medication records between the two cohorts: 53.5% CHD patients in UKB are taking lipid-lowering medications, while this number is only 1.78% in SHS. We further performed a sensitivity analysis by excluding all individuals taking lipid medications in both cohorts, and found that APOB switched to be a risk factor in the subset of UKB participants as well (effect size = 0.04, p-value = 4.1E-4), consistent with SHS results and prior studies^29^.

**Figure 3.**
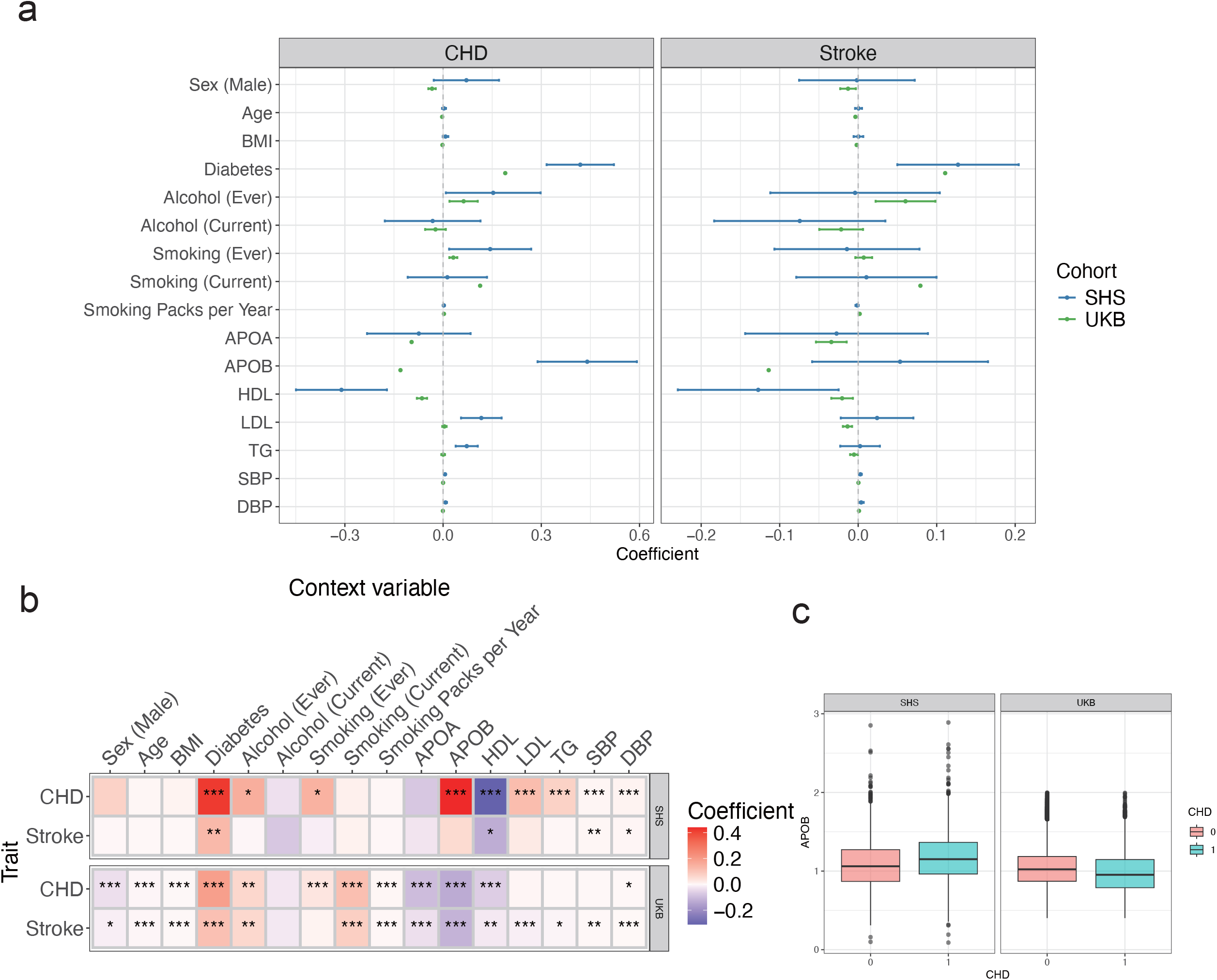
Univariate tests of association between cardiovascular traits and context variables. **a**. Forest plots of coefficients of context variables for coronary heart disease (CHD) and stroke in SHS (blue lines) and UKB (green lines). The x-axis represents the effect size of each context variable and error bars indicate 95% confidence intervals. **b**. Heatmap of coefficients of context variables for CHD and stroke in SHS (upper panel) and UKB (lower panel). Asterisks represent significance (p-value < 0.05(*)/0.01(**)/0.001(***)). **c**. Distribution of Apolipoprotein B (APOB) in CHD patients (CHD = 1) and healthy individuals (CHD = 0) in SHS and UKB for all of our study participants (before removing individuals taking statin).

Furthermore, differing from the associations with BP, current smoking significantly increased the risk for both cardiovascular events in UKB, and a history of smoking was also associated with increased CHD risk in both populations.

### Improved prediction adding context variables to PGS

Following the screening of significant context variables in training, we selected trait-specific context variables for SHS and UKB (**Table 2**) from trained prediction models, which were further applied to the testing set to quantify the improvement in prediction performance compared to genetics-only models (**Methods**). For BP traits, the average partial R^2^ in SHS and UKB were 2.05% and 1.80% (**Figure 4a, Supplementary Table 5**). DBP achieved the highest gain from adding context variables to PGS in both populations (SHS: R^2^ = 7.29% vs 4.56%, p-value = 1.1E-4; UKB: R^2^ = 20.47% vs 17.98%, p-value = 1.2E-58). For CHD and stroke, the benefit of incorporating context information varied between the two cohorts (**Figure 4b, Supplementary Table 6**). In SHS, adding N=9 context variables significantly increased the prediction AUC of CHD from 0.656 to 0.740 (p-value = 5.7E-12), and marginally improved AUC for stroke (AUC = 0.667 to 0.636, p-value = 0.06). As expected, given that most context variables were significantly associated with both cardiovascular outcomes in UKB, the AUC increased from 0.759 to 0.792 for CHD (p-value = 1.3E-25) and from 0.697 to 0.755 for stroke (p-value = 6.2E-16).

**Table 2.** Trait-specific selected significant context variables.

| Trait | Cohort | Selected context variables |
| --- | --- | --- |
| SBP | SHS | Diabetes, Smoking status, Smoking pack per year, APOA, APOB, HDL, TG |
|  | UKB | Diabetes, Alcohol status, Smoking status, APOA, APOB, HDL, LDL, TG |
| DBP | SHS | Diabetes, Smoking status, Smoking pack per year, APOA, APOB, HDL, TG |
|  | UKB | Alcohol status, Smoking status, Smoking pack per year, APOA, APOB, HDL, LDL, TG |
| PP | SHS | Diabetes, APOA, HDL |
|  | UKB | Diabetes, Alcohol status, Smoking status, Smoking pack per year, APOA, HDL, LDL, TG |
| CHD | SHS | Diabetes, Alcohol status, Smoking status, APOB, HDL, LDL, TG, SBP, DBP |
|  | UKB | Sex, Age, BMI, Diabetes, Alcohol status, Smoking status, Smoking pack per year, APOA, APOB, HDL, DBP |
| Stroke | SHS | Diabetes, HDL, SBP, DBP |
|  | UKB | Sex, Age, BMI, Diabetes, Alcohol status, Smoking status, Smoking pack per year, APOA, APOB, HDL, LDL, TG, SBP, DBP |

**Figure 4.**
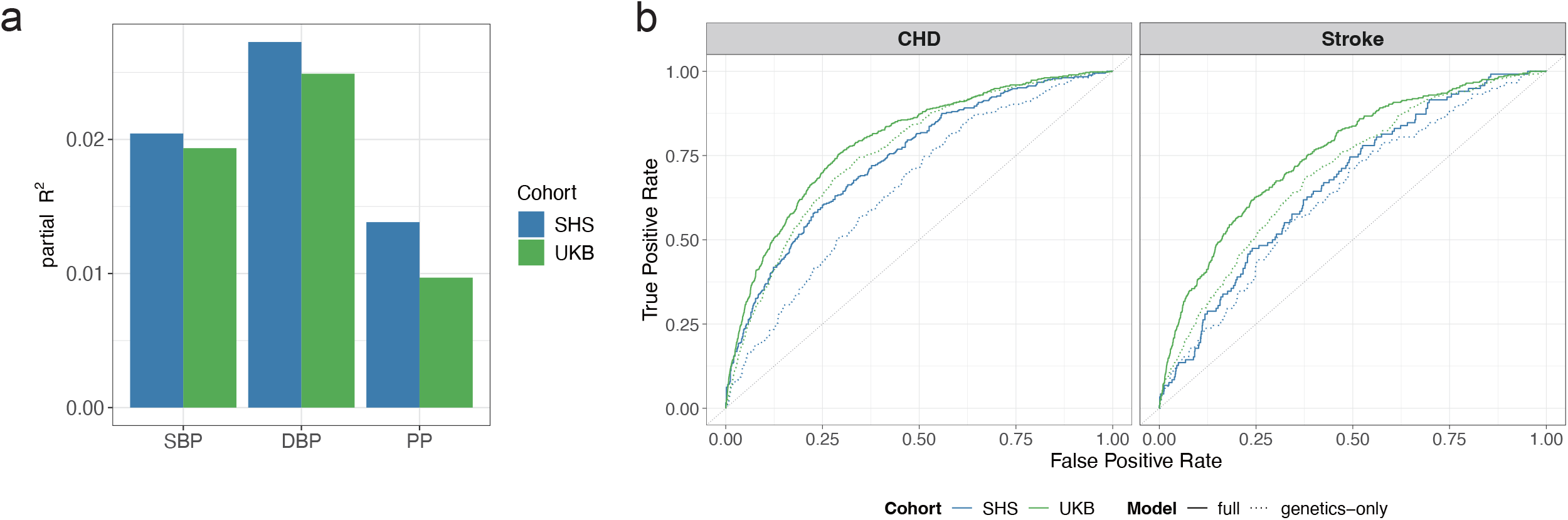
Improvement in prediction by integrating context variables. **a**. Bar chart showing the improvement in model fit (partial R^2^) for blood pressure traits (SBP, DBP, PP) when adding context variables and their interactions with PGS to a genetics-only base model. Blue bars represent SHS, and green bars represent UKB. **b**. Receiver Operating Characteristic (ROC) curves for CHD and stroke prediction models. Solid lines represent the full model (covariates + PGS + context variables + interactions), and dashed lines represent the “genetics-only” model (covariates + PGS). The integration of context variables significantly improved the Area Under the Curve (AUC) for both traits in both cohorts.

### Improved CHD risk prediction compared to an established model for American Indians

We further assessed the value of integrating PGS with the identified context variables (as well as their interactions) compared to an established CHD risk prediction model specifically for American Indians^28^. We note that such a model has already included some of our identified context variables as clinical predictors, such as SBP, HDL, LDL, diabetes, and smoking status. Using the same context variables, we showed a lower C-index to the original model (0.67 vs. 0.73 for female and 0.64 vs. 0.70 for male, for our model and established model, respectively), likely due to the difference in sample size (N = 2,158 vs. 4,372 in our model and the established model, respectively). Our GxC model, however, achieved significantly improved CHD prediction in our SHS cohort compared to the original publication^28^. The model’s C-index increased from 0.672 to 0.705 in females (LRT p-value = 3.9E-7) and from 0.644 to 0.675 in males (LRT p-value = 4.8E-5). A similar significant improvement was observed for the entire cohort without sex stratification (C-index: 0.66 to 0.684; LRT p-value = 1.2E-10).

## Discussion

In this study, we identified context variables that are associated with BP and cardiovascular diseases (specifically CHD and stroke) while accounting for PGS, and evaluated the utility of integrating these context variables with PGS to enhance risk prediction in both European and American Indian populations. Our findings highlight two critical advances in precision medicine. First, we demonstrated that incorporating context variables and their interactions with genetic risk significantly improves predictive performance beyond PGS alone. Second, by adding these GxC interaction terms to an established clinical risk model for American Indians, we achieved a significant improvement in model discrimination, suggesting tangible clinical utility for this underrepresented population.

We observed both shared and population-specific patterns in how context variables influence cardiometabolic risk. Consistent across both cohorts, diabetes status emerged as a robust risk factor, and HDL as a protective factor for cardiovascular outcomes. Interestingly, we observed the “smoking paradox” in both populations, where current smoking was associated with lower SBP and DBP^30,31^ but significantly increased the risk of CHD and stroke, likely due to other harmful effects. This underscores the complexity of context-specific effects, where a factor may appear beneficial for an intermediate phenotype (e.g., BP) while remaining detrimental to the ultimate disease endpoint through other endophenotypes or pathways. Additionally, we originally found that higher APOB levels were associated with increased CHD risk in SHS but appeared protective in UKB, but this spurious result in UKB may have been confounded by medication usage. The absence of this pattern in SHS suggests differences in medication usage patterns or baseline risk profiles between the cohorts during the study periods. The low lipid-lowering medication usage for individuals with CHD in SHS (1.78%) is in part a reflection of their limited access to healthcare services, emphasizing the importance for genetic epidemiological research in this population, and for other stakeholders to take relevant actions.

A major implication of our work is the potential to refine clinical risk assessment for American Indians. By integrating PGS with our identified context variables, we built an improved CHD risk prediction model compared to a previously established model for American Indians. This improvement is particularly notable given that the PGS weights were largely derived from European ancestry populations, which typically leads to attenuated performance in non-European ancestry individuals. Our results suggest that even with imperfectly transferrable PGS, the rigorous integration of lifestyle and clinical contexts can recover substantial predictive power, bridging part of the gap in equitable precision medicine.

Although showing promising clinical implications, our study also has limitations. First, the sample size disparity of PGS set between UKB (N ∼ 121K) and SHS (N ∼ 3K) results in differential statistical power, likely explaining why fewer context variables were statistically significant in SHS. Second, while we adjusted for antihypertensive and lipid lowering medications (but only for LDL), residual confounding may still influence gene-context interaction estimates, which has been proven in our sensitivity analysis for APOB. Finally, the context variables used here are a subset of potential environmental exposures; future work could explore broader social determinants of health and environmental exposures that may further modulate genetic risk in diverse populations.

In conclusion, our study validates the GxC approach for cardiometabolic risk prediction. By moving beyond a “genetics-only” or “environment-only” perspective, we demonstrate that modeling the interplay between inherited risk and modifiable context factors offers a more accurate and population-sensitive tool for identifying individuals at high risk for cardiovascular disease.

## Methods

### Study populations and outcome definition

SHS is a cohort of 4,549 tribally-enrolled American Indians, aged 45 to 74 at initial recruitment from communities in Arizona, Oklahoma, and the Dakotas^32^. From 1989 to 1999, the study conducted three clinical examinations (Phase I: 1989-1991; Phase II: 1993-1995; Phase III: 1998-1999), collecting comprehensive clinical, lifestyle, and medication data, along with fasting blood and urine samples. Incident events were obtained from surveillance, mortality, and medical records using validated protocols, with a follow-up to December 31st, 2022. Incident CHD was defined by fatal and nonfatal definite myocardial infarction (MI), definite CHD, ECG-evident definite MI, cardiac procedures including percutaneous transluminal coronary angioplasty or coronary artery bypass graft events. Incident stroke included fatal and nonfatal stroke events adjudicated by two neurologists based on the International Diagnostic Criteria. Study protocols were approved by the Indian Health Services Institutional Review Board (IRB), IRBs of all Institutions and by the participating Tribal Review Boards. This study followed the Strengthening the Reporting of Genetic Association Studies (STREGA) reporting guideline. Of the participants, 3,221 consented to and were selected for genetic research.

UKB is a large biomedical database containing detailed genetic and health information from half a million volunteering participants in the United Kingdom^33^. In this study, we incorporated 424,622 individuals of primarily European ancestry, determined in our previous study^14^. CHD was defined using ICD-10 and ICD-9 codes (**Supplementary Table 7**), while other outcome variables were identified using specific data fields within UKB (**Supplementary Table 7**).

### Data harmonization and list of context variables

We consider three measurements for BP traits: SBP, DBP and PP. To account for the effects of antihypertensive medication, BP readings were adjusted for individuals reporting medication use in both cohorts, by adding 15 mmHg to their SBP and 10 mmHg to their DBP following previous study adjustment^34^.

The following context variables were selected: age, sex, BMI, alcohol consumption and smoking status (Never, Ever, and Current), smoking packs per year, diabetes status, and five lipid levels – Apolipoprotein A (APOA), Apolipoprotein B (APOB), high-density lipoprotein (HDL), low-density lipoprotein (LDL), and triglyceride (TG). For individuals on lipid medications, we adjusted LDL by dividing directly assessed LDL by 0.7, as previously recommended^35^, and used the adjusted LDL for all following analyses. For CHD and stroke, we also added SBP and DBP as context variables.

### SHS genome-wide genotyping

For SHS, genome-wide genotyping was performed for 1,748,250 markers using the Illumina MEGA array. Following quality control and imputation^2^, a final dataset of 13,210,147 variants for 3,157 individuals was available for downstream analysis.

### UKB Genome-wide Association Study (GWAS)

As most studies from the PGS Catalog^36^ included UKB in PGS training, we were unable to directly apply pre-trained PGS models in UKB. Thus, the UKB cohort was partitioned into a GWAS set (N = 300,000) and a PGS set (N = 124,622) for PGS development. To prevent information leakage due to familial relatedness, all related individuals (kinship > 0.005) were assigned to the training set. GWAS for 3 BP traits and 2 cardiovascular diseases (CHD and stroke) were conducted in the training set using a linear mixed model implemented in REGENIE-3.1.3^37^. Phenotypes were first adjusted for age, age^2^, sex, BMI, recruitment center, and top 10 genetic principal components (PCs), and the resulting residuals were inverse-normal transformed prior to association testing.

### Polygenic score construction

PGS were constructed for five traits in the SHS entire cohort and the UKB PGS set. Given the smaller sample size of the SHS, we constructed PGS using weights from previously published large-scale studies. Specifically, weights for BP traits were derived from a GWAS of over 1 million European individuals^34^, weights for CHD were derived and refined in Europeans^38^, and weights for stroke were from a meta-analysis of European and East Asian populations^39^. For UKB, we used our internal GWAS to construct PGS by PRS-CS^40^ with default parameters.

### Context variables evaluation

To quantify the predictive value of context variables beyond genetic risk (as captured by PGS), we conducted 2-step evaluation by further splitting the individuals with PGS in both cohorts into training (SHS, N = 2,000; UKB, N = 95,000) and testing (SHS, N = 1,221; UKB, N = 29,622). The initial screening was conducted in the training set by univariate tests. First, we generated the residuals from a baseline regression model (linear for BP measurements, logistic for CHD and stroke) against age, age^2^, sex, BMI and recruitment center. Then we fit a genetic-only model for the trait residuals with the top 10 PCs plus the trait-specific PGS (i.e., residuals ∼ 10PCs + PGS). To assess each context variable individually, we then fitted a series of GxC models, each containing all terms from the genetics-only model plus one single context variable (C) and its interaction term with the PGS (i.e., residuals ∼ 10PCs + PGS + C + PGS*C). The contribution of each context variable was estimated by its effect size in the GxC models. Those context variables which showed significant contributions (p-value < 0.05) were carried forward to the final evaluation, which was performed in the held-out testing sets. Here we fit a full model including baseline covariates, top 10 PCs, the trait-specific PGS, all selected context variables, and their corresponding interaction terms with the PGS (i.e., outcome ∼ sex + age + age^2^ + BMI + recruitment center + 10PCs + PGS + C_1_ + … + C_k_ + PGS*C_1_ + … + PGS*C_k_, where k is the total number of significant context variables selected by previous univariate tests). The performance of the full model was compared against the genetics-only model (i.e., outcome ∼ sex + age + age^2^ + BMI + recruitment center + 10PCs + PGS) using log-likelihood ratio tests (LRT). Performance improvement was assessed by the difference of the adjusted R^2^ (for BP traits) or AUC (for CHD and stroke) between the full and the genetic-only model.

### Comparison with an established CHD risk prediction model for American Indians

To determine if genetic information plus the context factors improve incident CHD risk prediction, we expanded upon a previously established clinical risk model specifically for American Indians, developed also in SHS^28^. We fitted two nested Cox proportional hazards (CoxPH) models on the SHS entire cohort after removing missingness. The baseline model included only the established clinical predictors from the prior study. The full model incorporated these clinical variables along with the PGS, significant context factors, and their interaction terms. We assessed the improvement in model fit using Harrell’s C-index and LRT. This analysis was performed both with and without sex-stratification, concordant with the prior study.

## Acknowledgments

We want to express our gratitude to the many SHS and other participants, as well as the Tribal governments that generously offered their time and effort to this cause.

## Data availability

The data were collected, analyzed, and reported under agreements made with the sovereign tribal nations that have partnered in this research, which preclude routine modes of data sharing. Requests to access the dataset from qualified researchers trained in human subject confidentiality protocols may be sent to the Strong Heart Study Coordinating Center at https://strongheartstudy.org/. Requests will be reviewed by tribal research partners before data may be released. This policy is consistent with the “NIH Policy for Data Management and Sharing: Responsible Management and Sharing of American Indian/Alaska Native Participant Data.”^41^

