## Supplementary Figures for "Bridging the Genomic Equity Gap with Context-Enhanced Risk Stratification in American Indians: the Strong Heart Study"

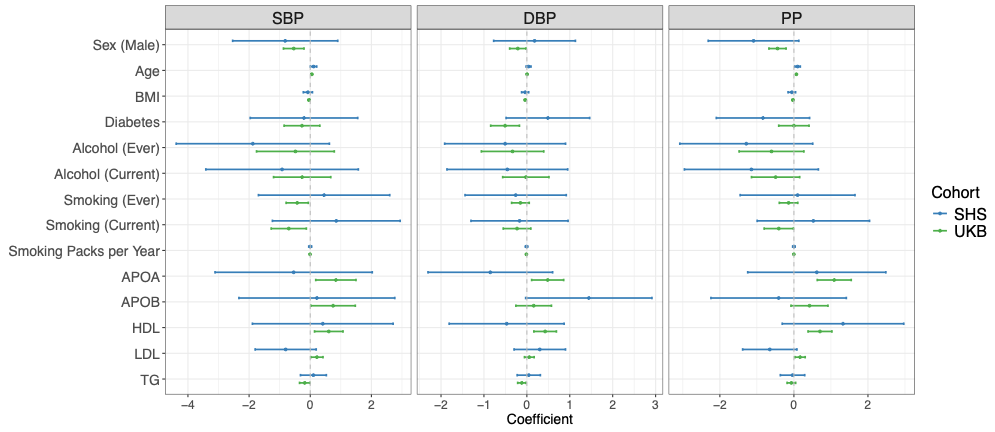

**Supplementary Figure 1.** Forest plots of univariate test coefficients of context variables*PGS for systolic blood pressure (SBP), diastolic blood pressure (DBP), and pulse pressure (PP) in SHS (blue lines) and UKB (green lines). The x-axis represents the effect size of each context variable and error bars indicate 95% confidence intervals.

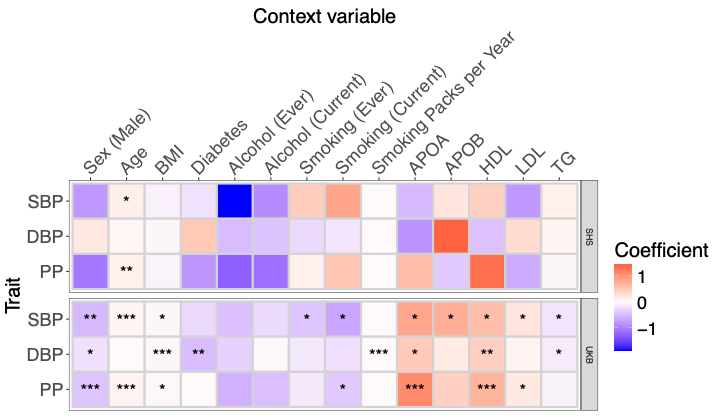

**Supplementary Figure 2.** Heatmap of univariate tests coefficients of context variables*PGS for SBP, DBP and PP in SHS (upper panel) and UKB (lower panel). Asterisks represent significance (p-value < 0.05(*)/0.01(**)/0.001(***)).

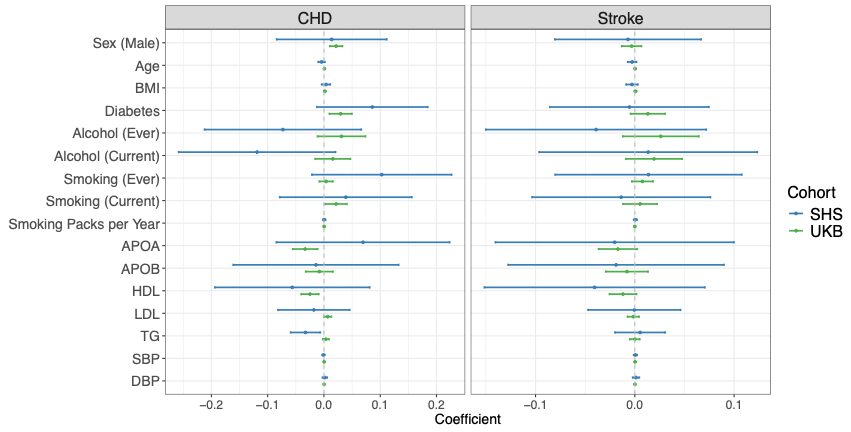

**Supplementary Figure 3.** Forest plots of univariate test coefficients of context variables*PGS for coronary heart disease (CHD) and stroke in SHS (blue lines) and UKB (green lines). The x-axis represents the effect size of each context variable and error bars indicate 95% confidence intervals.

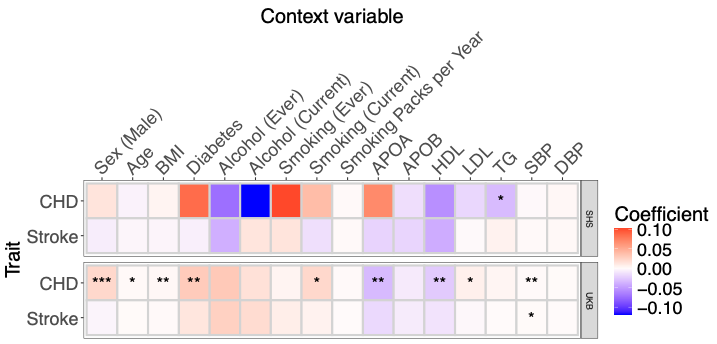

**Supplementary Figure 4.** Heatmap of univariate tests coefficients of context variables*PGS for coronary heart disease (CHD) and stroke in SHS (upper panel) and UKB (lower panel). Asterisks represent significance (p-value < 0.05(*)/0.01(**)/0.001(***)).
